# Neonatal AKI profile using KDIGO guidelines: A cohort study in tertiary care hospital ICU of Lahore, Pakistan

**DOI:** 10.1101/2022.03.14.22272344

**Authors:** Rafia Gul, Zahid Anwar, Mehmood Sheikh, Ayesha Salamat, Samer Iqbal, Furqan Saleem

## Abstract

**Background:** AKI is witnessed in sick neonates and is associated with poor outcomes. Our cohort represents the profile of neonates who were diagnosed with AKI using KDIGO guidelines during intensive care unit stay.

**Methodology:** A cohort study was conducted in the NICU of FMH from June 2019 to May 2021. Data were collected on standardized proforma. Serum creatinine was measured within 24 hours after enrollment in the study by cytometric analysis using the C311 Rosch machine and subsequently after 24 to 48 hours. Data analysis was done using SPSS v 20.0. All continuous variables were not normally distributed and were expressed as the median and interquartile range (IQR). Categorical variables were analyzed by proportional differences with either the Pearson chi-square test or Fisher’s exact tests. A multinomial logistic regression model was used to explore the independent risk factors of AKI. Time to the event (death) and survival curves for the cohort were plotted by using Cox proportional hazard model.

**Results:** AKI occurred in 473 (37.6%) of neonates and 15.7%, 16.3% and 5.6% had stage 1, 2 and 3 respectively. The outborn birth (p 0.000, AOR 3.987, 95%CI 2.564 – 6.200), birth asphyxia (p 0.000, AOR 3.567, 95%CI 2.093 – 6.080), inotropic agent (p 0.000, AOR 2.060, 95%CI 1.436 – 2.957), antenatal steroids (p 0.002, AOR 1.721, 95%CI 1.213 – 2.443), central lines (p 0.005, AOR 1.630, 95%CI 1.155 – 2.298), IVH/ICH/DIC (p 0.009, AOR1.580, 95%CI 1.119 – 2.231) and NEC (p 0.054, AOR 1.747, 95%CI 0.990 – 3.083) were independently associated with AKI. Protective factors of neonatal AKI were normal sodium levels, maternal diabetes mellitus as well Hb>10.5 mg/dl. Duration of stay (7 vs 9 days) and mortality rates (3.9% vs16.5%) were significantly higher in neonates with AKI (p <0.001).

**Conclusion:** About one-third of critically sick neonates had AKI. Significant risk factors for AKI were outborn birth (298%), birth asphyxia (256%), inotropic agents (106%) %, NEC 74.7%, antenatal steroids 72%, central lines 63% and IVH/ICH/DIC 58%. AKI prolongs the duration of stay and reduces the survival of sick neonates.

## Introduction

Acute kidney injury (AKI) is a condition characterized by an acute decline in kidney functions affecting fluid and electrolyte homeostasis and elimination of waste products [1]. Neonate are at increased risk of developing acute kidney injury because of relatively immature kidneys and low glomerular filtration rate [2,3]. In neonates, oliguria (urine output < 0.5 – 1.0 ml/kg/hr) may or may not be present in case of AKI [4]. renal failure can present with or without.^4^ AKI has been reported in every third neonate admitted in neonatal ICU [5].

Multiple maternal and neonatal features act as risk factors for neonatal AKI. Maternal risk factors include pre-labor rupture of membranes (PROM), antepartum hemorrhage (APH), hypertension, antenatal steroids, and drug intake like NSAIDs during pregnancy. While neonatal factors include birth asphyxia, prematurity, congenital anomalies of kidney & urinary tract (CAKUT), hemodynamic instability requiring inotropic support, sepsis, necrotizing enterocolitis (NEC), patent ductus arteriosus (PDA), nephrotoxic medications (like vancomycin, and aminoglycosides), coagulopathy, umbilical line placement [4,6].

Valuation of the severity, impact, and outcome of AKI in neonates has always been challenging. Literature review shows that different models like AKIN (Acute Kidney Injury Network) and RIFLE (Risk, Injury, and Failure; and Loss; and End-stage kidney disease) have been used for the diagnosis and severity categorization of AKI in neonates but none is universally accepted. The KDIGO (Kidney disease: improved global outcome) guidelines modified for neonates have been recently introduced for diagnosing and staging AKI [7].

International, as well as national statistics regarding neonatal AKI using KDIGO guidelines, are scarce. To fill in this gap, studies should be conducted to authenticate this guideline. Our study aims to determine the neonatal AKI profile including prevalence, possible etiological factors, staging of acute kidney injury, and outcome using KDIGO guidelines in tertiary care hospital ICU.

## Material and methods

A prospective cohort study was conducted after ethical approval from the Institutional Review Board (IRB). The study period spans over 2 years from June 2019 to May 2021 in the department of neonatology, Fatima Memorial Hospital, Lahore. Our unit is Level – 3 with 26 bedded intensive care beds and one of the largest NICU in the province. We enrolled all neonates who were admitted to NICU during the study period after seeking consent from parents or guardians. However, all those neonates having major anomalies incompatible with life, congenital renal disorders, who did not survive until 72 hours of postnatal life, or parents who refused to grant permission to participate in the study were excluded. Serum creatinine was measured by cytometric analysis using the C311 Rosch machine.

## Data Collection

All neonates admitted to our NICU, fulfilling inclusion criteria with any clinical diagnosis were enrolled in the study and managed as per departmental protocols. All admitted neonates were assessed for urine output (ml/kg/hour) and serum creatinine.

The serum creatinine level of all neonates was measured within 24hours after enrollment in the study and was taken as a baseline. Further serum creatinine levels were checked after every 24 to 48 hours in case-specific conditions. In the current study, serum creatinine level was used to classify the degree of AKI following KDIGO guidelines. KDIGO diagnosees and classifies AKI in a tiered fashion by using urine output and serum creatinine values as follows [7]. However, AKI was never labeled in any neonate within the first 72 hours of life.

- Stage 0: No change in S/Cr or rise <0.3 mg/dl or urine output ≥0.5 ml/kg/hour
- Stage 1: S/Cr rise ≥0.3 mg/dl within 48 hours or S/Cr rise ≥1.5 – 1.9 × baseline previous S/Cr or <0.5 ml/kg/hour for 6 – 12 hours
- Stage 2: S/Cr rise ≥2.0 – 2.9 times baseline previous S/Cr or <0.5 ml/kg/hour for ≥12 hours
- Stage 3: S/Cr rise ≥3.0 times baseline previous S/Cr or S/Cr ≥2.5 mg/dl (<10 ml/min/1.73m^2^) or candidate for renal replacement therapy or <0.3 ml/kg/hour for ≥24 hours or anuria for ≥12 hours.

Schwartz’s formula [8] was used to calculate GFR as follows

eGFR = k x length (cm) /S. creatinine (mg/dl)

k = constant 0.33 for low birth weight and 0.45 for term babies

The highest serum creatinine of neonates was used to calculate GFR.

Maternal demographic data including diabetes mellitus, hypertensive disorders, antepartum hemorrhage (APH), parity, maternal anemia, smoking, addiction, and antenatal steroids during pregnancy was collected. Neonatal data including demographic [gestation age, weight, length, gender, delivery mode, and age of neonate], clinical characteristics [mechanical ventilation (noninvasive/invasive), central line (umbilical catheterization and peripherally inserted cardiac catheter), use of medications such as inotropic agents and nephrotoxic medications (vancomycin, aminoglycosides)] and comorbid conditions [sepsis (leukocytosis or leukopenia or ANC<150 along with raised CRP and platelets <100 and or positive blood culture), presence of hemodynamically significant patent ductus arteriosus (hsPDA), intraventricular hemorrhage (IVH), intracranial hemorrhage (ICH), disseminated intravascular coagulopathy (DIC) and necrotizing enterocolitis (NEC) (modified Bell’s stage II or III)].

All neonates were monitored for the duration of hospital stay and their outcome in form of discharge or expiry.

### Data Analysis

Data was analyzed using Statistical Package for the Social Sciences (SPSS 20.0) package. Descriptive statistics and tests of significance were calculated for all the variables. The Shapiro Wilk test was used to assess the normality of the distribution of investigated continuous parameters (length, weight, gestation age, and glomerular filtration rate GFR). All continuous variables were not normally distributed and were expressed as the median and interquartile range (IQR). Categorical variables were analyzed by proportional differences with either the Pearson chi-square test or Fisher’s exact tests. The z-test was applied to compare column proportions and p-values were adjusted using the Bonferroni method. The level of significance was considered with p <0.05. To identify independent risk factors for AKI, demographic data and clinical characteristics were compared between neonates with and without AKI. A multinomial logistic regression model was used to explore the independent risk factors of AKI. Time to the event (death) and survival curves for the cohort were plotted by using Cox proportional hazard model.

## Results

During the study period, 1680 neonates were admitted to our neonatal intensive care units, out of which 239 were excluded. Of 1364 neonates, 1258 (92.2%) had finally complete data for final analysis (figure 1).

**Figure 1:**
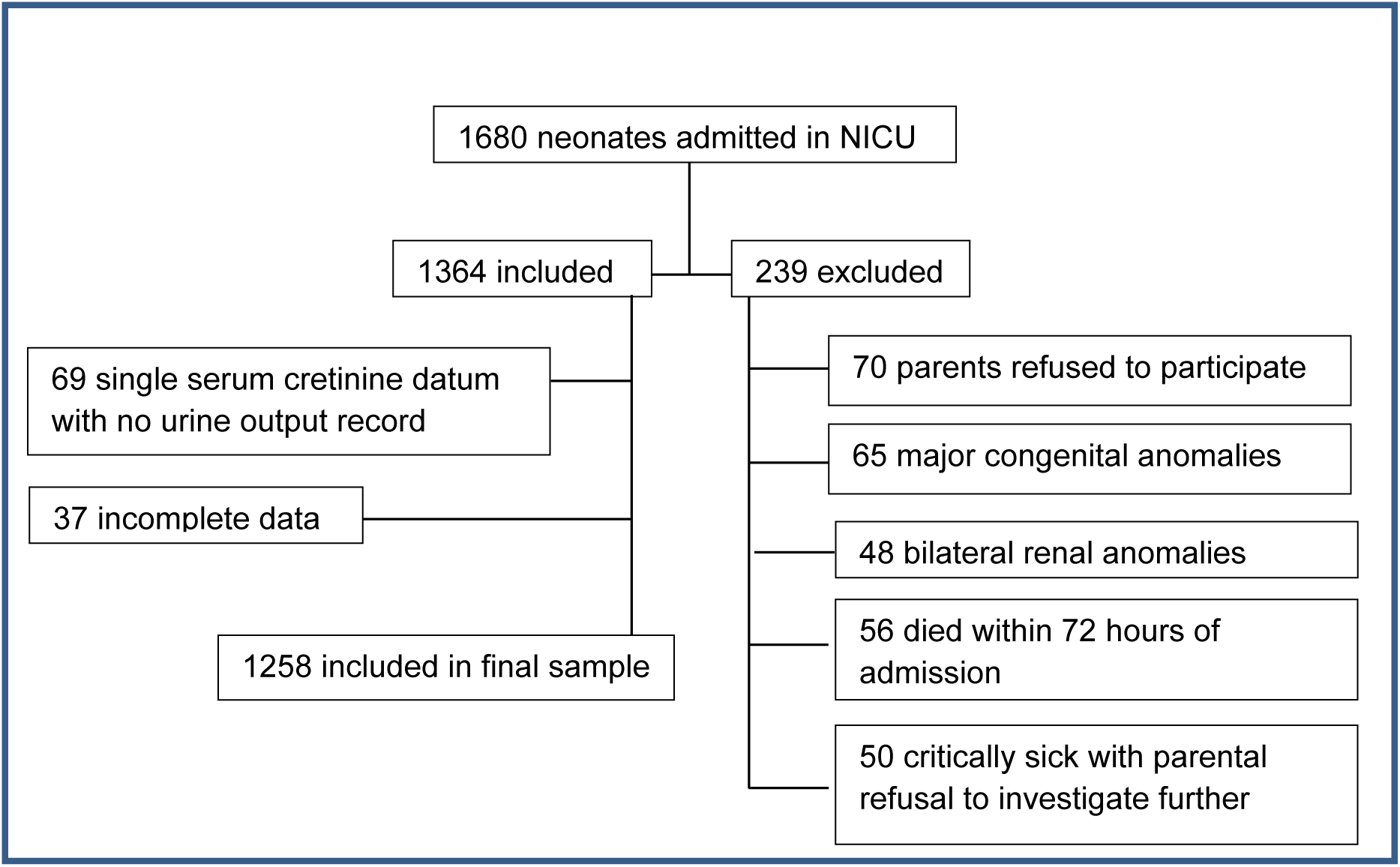
Flow chart of selected patients

Of these 1258 neonates 473 (37.6%) had AKI and among these 197 (15.7%) had stage 1 AKI, 205 (16.3%) stage 2 and only 71 (5.6%) had stage 3 AKI. The gestational age-based stratification of the cohort shows that 39 (3.1%) neonates were extremely preterm (gestation age < 28 weeks), 878 (69.8%) of gestation age 28 to 36+6 weeks, and 341(27.1%) were of gestation age 37 - 42 weeks. Neonates of gestation age 28 weeks or less had proportionally more incidence of AKI p<0.001. There were 68 neonates of weight <1kg, 350 of 1.1 – 1.5kg, 480 of 1.6 – 2.5kg and 350 of >2.5kg. In neonates of <1 kg, 1.1 to 1.5 kg, 1.6 to 2.5kg and > 2.5 kg AKI was documented in 57.4 %, 38.3%, 36.7% and 34.3% respectively. Maternal and neonatal characteristics with and without AKI have been shown in table 1.

**Table 1:**
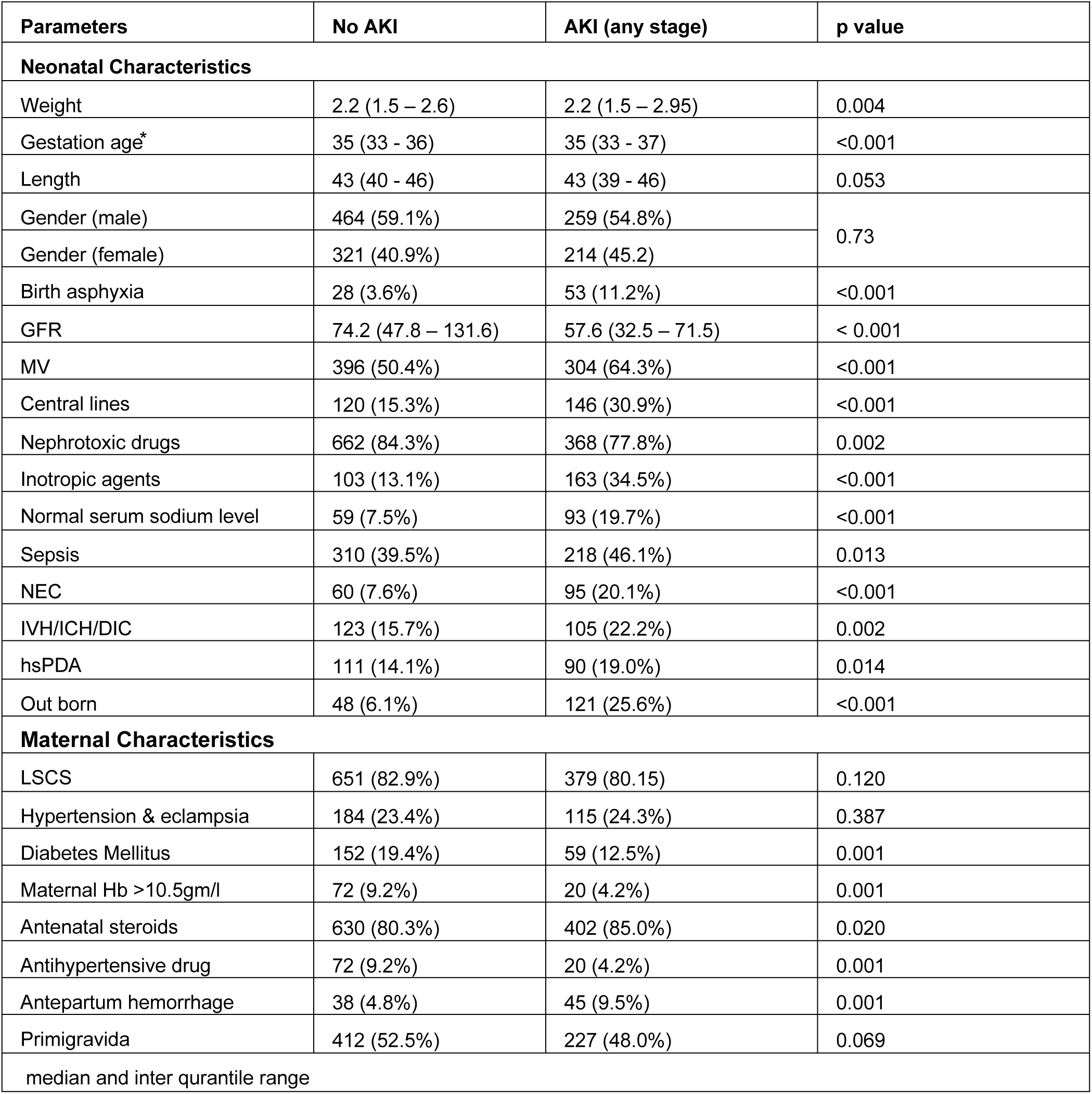
Maternal and neonatal characteristics with and without AKI

Maternal diabetes mellitus, maternal Hb>10.5 mg/dl and antepartum hemorrhage along with the use of antihypertensive drugs and antenatal steroids were risk factors to neonatal AKI. Neonatal gender, birth order, cesarean section as the mode of delivery, and maternal history of hypertension or preeclampsia were statistically insignificant as a risk factor of neonatal AKI.

The outborn neonates had a 298% high risk of developing AKI as compared to inborn. Birth asphyxia increased the risk of developing AKI by 256% followed by inotropic agent 106%, NEC 74.7%, antenatal steroids 72%, central lines 63%, IVH/ICH/DIC 58%, sepsis 6.1%, hsPDA 5.6%, and mechanical ventilation 1%. Neonates with normal sodium levels have 48.5% less risk of developing AKI. Maternal diabetes mellitus as well Hb>10.5 were protective factors for neonatal AKI by 56.4% and 46.5% respectively. These all factors were statistically significant (table 2).

**Table 2:**
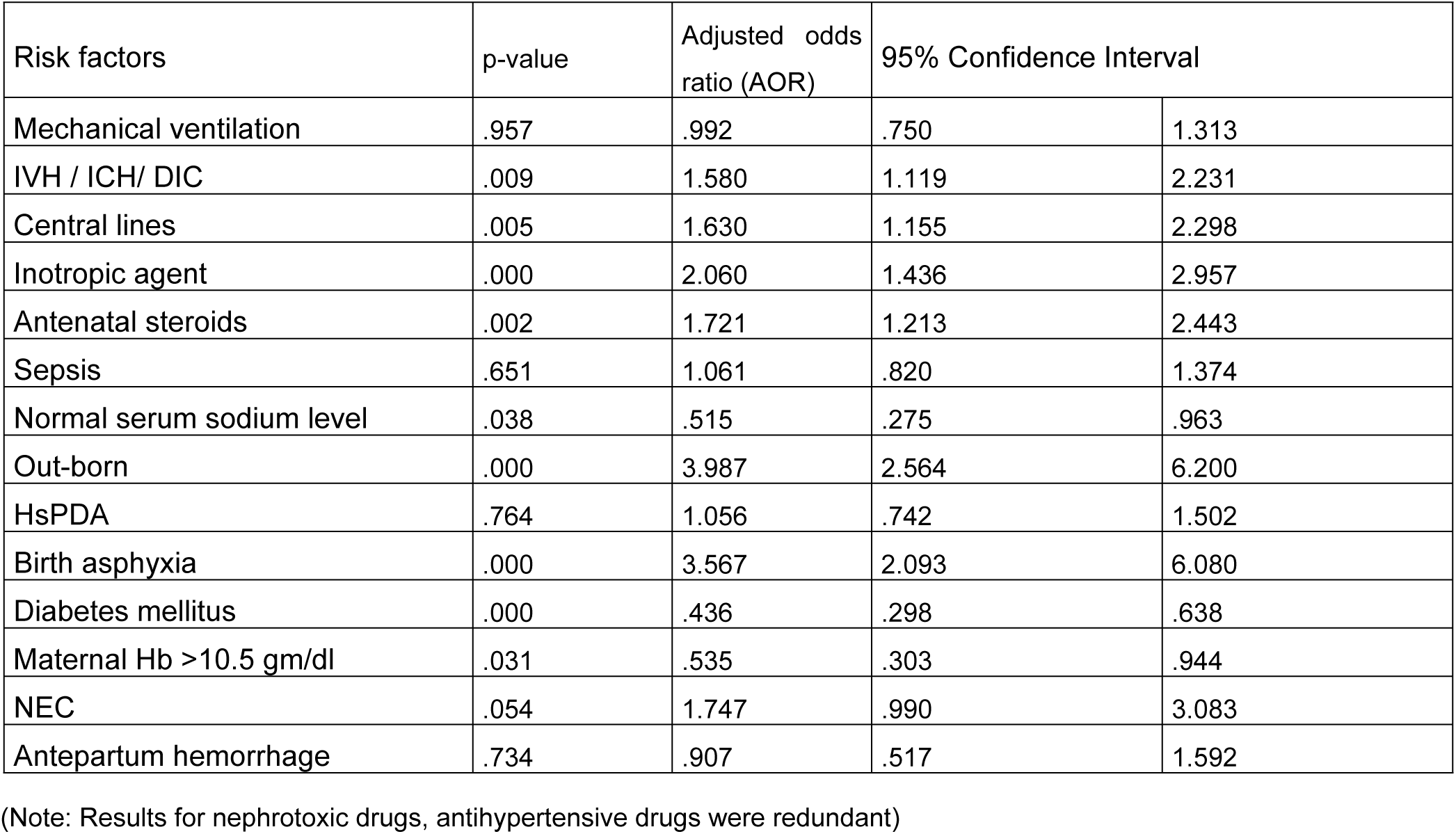
Risk factors for AKI

| Risk factors | p-value | Adjusted odds ratio (AOR) | 95% Confidence Interval |  |
| --- | --- | --- | --- | --- |
| Mechanical ventilation | .957 | .992 | .750 | 1.313 |
| IVH / ICH/ DIC | .009 | 1.580 | 1.119 | 2.231 |
| Central lines | .005 | 1.630 | 1.155 | 2.298 |
| Inotropic agent | .000 | 2.060 | 1.436 | 2.957 |
| Antenatal steroids | .002 | 1.721 | 1.213 | 2.443 |
| Sepsis | .651 | 1.061 | .820 | 1.374 |
| Normal serum sodium level | .038 | .515 | .275 | .963 |
| Out-born | .000 | 3.987 | 2.564 | 6.200 |
| HsPDA | .764 | 1.056 | .742 | 1.502 |
| Birth asphyxia | .000 | 3.567 | 2.093 | 6.080 |
| Diabetes mellitus | .000 | .436 | .298 | .638 |
| Maternal Hb >10.5 gm/dl | .031 | .535 | .303 | .944 |
| NEC | .054 | 1.747 | .990 | 3.083 |
| Antepartum hemorrhage | .734 | .907 | .517 | 1.592 |

(Note: Results for nephrotoxic drugs, antihypertensive drugs were redundant)

Mortality rates were higher in neonates with AKI as compared to the non-AKI group. Of 473 neonates with AKI, 16.5% expired (p <0.001). Among all neonates with acute kidney injury, neonates with AKI stage 3 had higher mortality rates (42.3%) as compared to stage 1 (8.1%) and stage 2 (15.6%) respectively. Similarly, length of stay was directly related to the severity of AKI. Length of stay was lengthier in neonates with acute kidney injury (table 3).

**Table 3:**
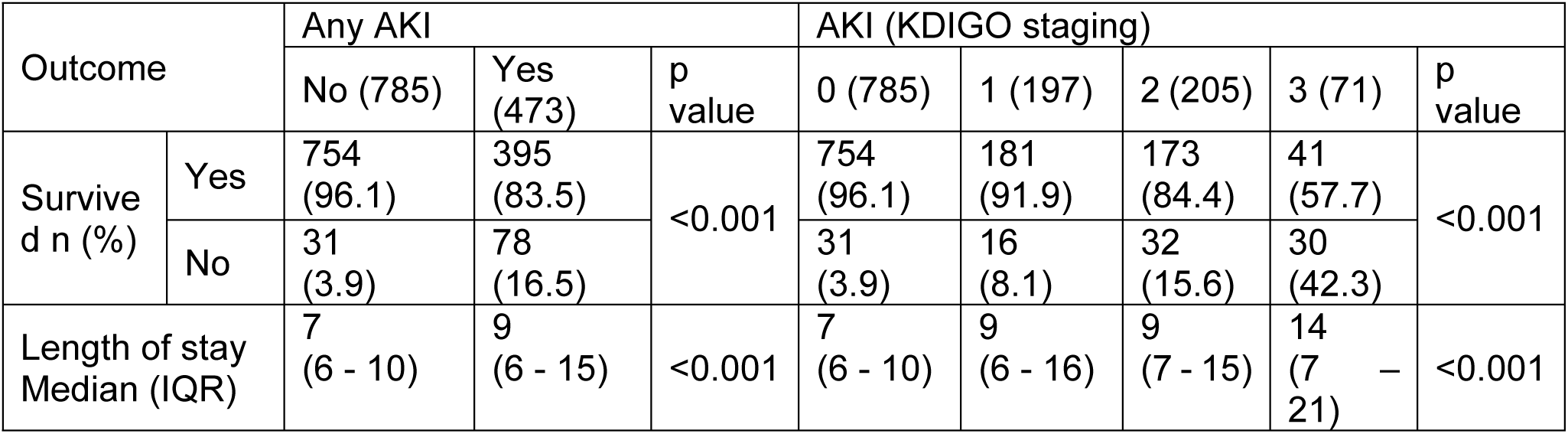
Duration of stay and clinical outcomes by AKI status

| Outcome |  | Any AKI |  |  | AKI (KDIGO staging) |  |  |  |  |
| --- | --- | --- | --- | --- | --- | --- | --- | --- | --- |
|  |  | No (785) | Yes (473) | p value | 0 (785) | 1 (197) | 2 (205) | 3 (71) | p value |
| Survived n (%) | Yes | 754 (96.1) | 395 (83.5) | <0.001 | 754 (96.1) | 181 (91.9) | 173 (84.4) | 41 (57.7) | <0.001 |
|  | No | 31 (3.9) | 78 (16.5) |  | 31 (3.9) | 16 (8.1) | 32 (15.6) | 30 (42.3) |  |
| Length of stay Median (IQR) |  | 7 (6 - 10) | 9 (6 - 15) | <0.001 | 7 (6 - 10) | 9 (6 - 16) | 9 (7 - 15) | 14 (7 - 21) | <0.001 |

Survival curves for the cohort were plotted by using Cox proportional hazard model (figure 2). Neonates with AKI had compromised survival outcomes and most of these expired within 30 days of hospital stay (figure 2).

**Figure 2:**
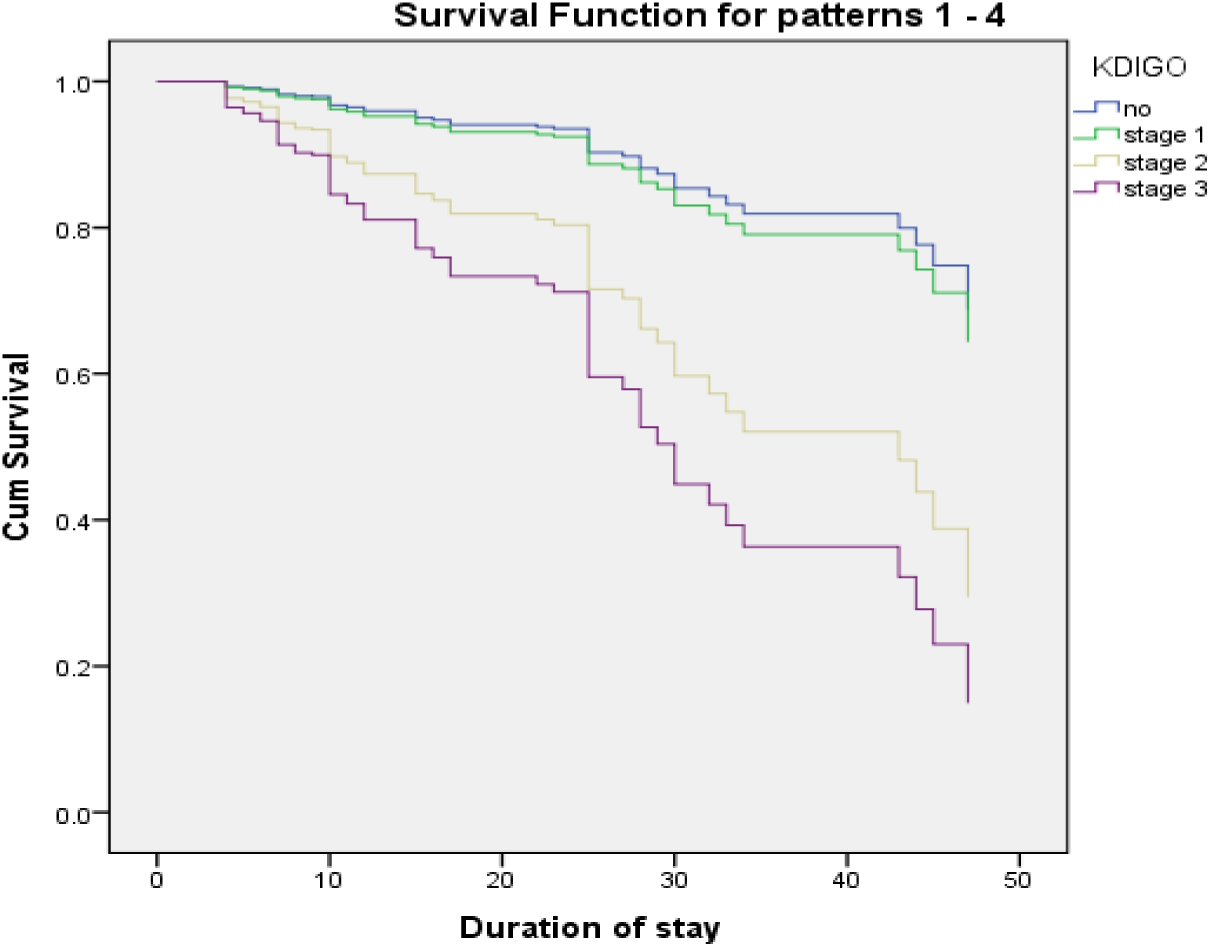
Survival curves

## Discussion

AKI is independently associated with the clinically significant outcome as it appears to be a critical component in the disease process. Only limited data is available regarding the diagnosis of AKI in neonates while using recently introduced KDIGO guidelines specially modified for neonates.

AKI is common in sick neonates admitted to NICU. Charlton and Jetton have reported the incidence of AKI in 21% and 30% of sick neonates admitted in NICU respectively [5,9]. More than one-third of neonates of our cohort had AKI. The similarity in findings is attributable because of resemblance in study settings and all gestation ages were recruited. The other hand, Alora reported that 11.7% of term neonates with perinatal asphyxia developed AKI [10]. Mazaheri documented that 9.7% of neonates developed AKI [11]. These differences are because in this study serum creatinine >1.5mg/dl as diagnostic criteria.

No gestational age is exempted to AKI. However, neonates of extreme gestation age <28 and >36 weeks are more vulnerable as compared to neonates in the middle group of gestation age 28 – 36 weeks [5,9,12]. In preterm and low birth weight neoantes prevelnce of AKI has been reprted as 56% by Lee, 63.3% Lei and 56.1% Shalaby respectively [13-15]. Our prterms of <28 weeks had almost similar AKI incidence at 43.6%. Understandably, immature kidneys with immature nephrogenesis are quite vulnerable to AKI especially in presence of other risk factors [16]. According to our cohort, the AKI rate in neonates born at 37 weeks and above is consistent with those in other studies of term neonates [17,18]. The potential justifications of this trend could be the ongoing process of nephrogenesis and renal maturation in these preterm neonates which is further compromised in presence of risk factors [19].

Literature review reveals that out-born birth was a unique risk factor for AKI in neonates. All sick neonates who require transfer due to any medical or surgical reason are at high risk of developing AKI [5,9,20]. Gohiya has reported a significantly higher incidence (p=0.02) of AKI in outborn neonates [21]. Similarly, our study shows that all neonates who were out-born and referred to our tertiary level care nursey had 4 times higher odds of AKI as compared to inborn neonates. One possible explanation could be an association of comorbid conditions leading to renal damage and consequently requiring transfer to tertiary level care for ideal management.

Several studies have shown that the severity of renal failure is directly related to the severity of asphyxia. The incidence ranges from 9.1% to 56%with moderate to severe asphyxia [22]. According to Gallo, Momtaz, and Elmas 70%, 4%, and 20 % of the study population with birth asphyxia developed AKI respectively [23-25]. Birth asphyxia is attributed to a decrease in renal blood flow and an increase in renal vascular resistance which is contributed by acidosis, high level of angiotensin II, prostaglandins, nitric oxide, and catecholamine [1,19,26]. Our cohort shows that neonates with birth asphyxia had a 256% higher risk of developing AKI. Variable association is attributed to difference in diagnostic citeria of AKI.

Neonates requiring inotropic support have a higher incidence of AKI as has been reported in multiple studies.[6,10,11,25,27]. All of our neonates who required inotropic agents had double the odds of AKI. It has been speculated that shock requiring inotropic agents as per se is a promotor of AKI as it alters renal blood flow and starts a cascade of renal damage by vasoconstriction [10].

NEC potentiates acute kidney insult. Hu et al have shown that risk of developing AKI increases 6 times in prence of AKI [28]. Our study shows that NEC increases the risk of developing AKI by 74.7%. This association is attributed to an inflammatory cascade which is a hallmark of NEC that leads to a disturbance at microcirculation level leading to progressive afferent arteriolar vasoconstriction and hence continuous ultrafiltrate loss [29].

According to Garg et al, all neonates whose mothers have received antenatal steroids are at higher risk (AOR 3.9, 95%CI 1.1 - 8.9) of developing AKI [20,30]. All neonates in our study has 72% higher risk of AKI. However, Ustun has reported the protective role of antenatal steroids regarding kidney injury [31]. The use of antenatal steroids for fetal lung maturation is common practice as part of international Obstetrics and Gynecology guidelines. Non-human models have proven that they affect fetal neohrogenesis by affecting nephron branching arrest and premature maturation [32].

Central catchers are lifelines for neonates during critical periods. Elmas et al [25] have observed a significant incidence of AKI in neonates having a central venous catheter (p 0.001) and umbilical catheter (p 0.002). AlGadeeb has documented that the presence of central lines, as well as umbilical arterial catheter, are significantly associated with AKI [27]. In our cohort, all neonates with central lines had a significantly higher incidence of AKI. One possible explanation could be combined damaging effect of central lines in prence of other risk factors.

AlGadeeb and Marciniak have reported intraventricular hemorrhage as a significant risk factor for neonatal acute kidney injury (p <0.001, AOR 2.605, 95%CI 1.465 - 4.631) and (OR 2.38, 95%CI 1.46 - 3.87) respectively [27,33]. DIC has been reported as a significant risk factor for neonatal acute kidney injury by Nickavar. ^6^ Our study highlights the association between intraventricular hemorrhage and disseminated intravascular coagulation with acute renal insult. Both DIC and IVH lead to a decrease in effective circulatory volume resulting in renal hypo-perfusion and failure [3].

Maternal diabetes mellitus does not appear to be an additional risk factor for neonatal renal insult [5,10,13]. Cappuccini et al have demonstrated that hyperglycemia has a negative impact on developing fetal kidneys resulting in reduced nephrogenesis, tubular integration, and renal failure [34]. However, Aisa et al have found that neonates of mothers who maintained strict glycemic control during pregnancy and fulfilled the other criteria of the GDM management program showed no differences compared to the normoglycemic group [35]. In our study, maternal diabetes mellitus rather appears to have a protective role against neonatal AKI. One possible explanation might be that during the antenatal period, these mothers have good glycemic control, vigilant monitoring, and regular checkup hence optimal fetal nephrogenesis.

The optimal fetal growth and development are coupled with optimal maternal health status. Maternal anemia is associated with adverse neonatal outcomes [5,9,12]. Our study population showed that all neonates of non-anemic mothers were protected from acute kidney injury. One possible explanation could be the impact of maternal hemoglobin status on fetal metabolic signaling pathway, hence affecting “ fetal programming” [36].

Dysnatremia either hypo or hypernatremia places neonates at risk of developing AKI. Hyponatremia was more prevalent in neonates with AKI as compared to neonates without AKI [37]. Hypernatremia has been reported as an important risk factor for AKI by Hamsa and Farhad [37-39]. However, Basalely has reported that dysnatremia was not associated with AKI [39]. Neonates with normal sodium levels have 48.5% less risk of developing AKI. Dysnatremia is associated with volume depletion and shock. Hence, it leads to decreased renal blood flow resulting in renal vasoconstriction, decreased GFR, and ultimately causes renal failure [2].

Mechanical ventilation, hsPDA, and sepsis are considered as significant risk factors for neonatal AKI [1,19,23-28]. Our study shows that these factors add risk to renal insult but are statistically insignificant. One possible explanation could be the homogeneous presence of these factors in all neonates of the cohort. Vancomycin and amikacin are considered to be nephrotoxic drugs and cause AKI [1,19,32]. However meta-analysis by Hu et all shows that a combination of these medications did not aggravate harmful effects to kidneys [28]. Similar results have been reflected in our study that show that these drugs have a negligible contribution to AKI.

The supporting data from other studies highlight that AKI is associated with poor outcomes [1-13]. Jetton et al analysis shows that neonates with AKI have 8.8 times longer duration of stay and 4.6 times higher mortality as compared to the non-AKI group [5]. Comparing different stages of AKI, the higher the order of renal failure poor is the outcome [11,20,21,26,27]. Irrespective of severity, all neonates in our cohort with AKI had significantly longer length of stay and higher odds of death. AKI in presence of other comorbid conditions requires medical treatment prolongs hospital stay and is responsible for the poor neonatal outcome.

## Conclusion

About one-third of critically sick neonates developed AKI. Outborn birth (298%), birth asphyxia (256%), inotropic agents (106%) were significant risk factors for AKI. Other risk factors include, antenatal steroids, central lines, IVH/ICH/DIC, and NEC. AKI prolongs the duration of stay and reduces the survival of sick neonates.

## Data Availability

Data will be available after acceptance of manuscript

## LIMITATIONS

There are several limitations to this study. First of all only serum creatinine level was used to diagnose and stage renal failure and urine output was not incorporated. Urine output monitoring by catheterization was never planned to avoid catheter-associated urinary tract infection. Pamper weight method was also not selected because of the frequent presence of stool with urine in pampering. Second, the long-term outcome was neither planned nor monitored. Third, the higher incidence of AKI would have been expected if it had included all neonates with bilateral renal anomalies, major congenital anomalies, and criticality sick neonates who were first enrolled then excluded due to parental refusal.

